# Validation of the Care Partner Stress Scale in the CAN-PROTECT Study

**DOI:** 10.1101/2025.09.04.25335106

**Authors:** Daniella Vellone, Dylan X. Guan, Jasper F. E. Crockford, Ibadat Warring, Clive Ballard, Byron Creese, Anne Corbett, Ellie Pickering, Pamela Roach, Eric E. Smith, Zahinoor Ismail

## Abstract

**Background:** Family and friend care partners play a vital role in supporting individuals with neurocognitive disorders, such as Alzheimer disease and related dementias. Care partners are often uncompensated and face unique, multifaceted challenges that contribute to overall stress. The Care Partner Stress Scale (CPSS) was developed to assess caregiver stress across seven domains: cognition, behaviour, function; unmet needs and emotional impact; work and financial strain; family and interpersonal conflict; and situational perception.

**Objective:** To evaluate psychometric properties of the CPSS in care partners of individuals with neurocognitive or neurodegenerative diseases of aging.

**Methods:** The CPSS was completed by 116 (87.1% female, age=62.11 years) care partner participants in CAN-PROTECT. Participants completed measures of depression, anxiety, quality of life, function, loneliness, and life satisfaction and engagement. We assessed internal consistency, item-total correlations, convergent and discriminant validity, and floor/ceiling effects.

**Results:** The CPSS demonstrated excellent internal consistency (α=0.96, 95% CI: 0.94-0.97), with all item-total correlations >0.30. Higher scores were associated with greater depression (*b*=2.73, 95%CI [0.82, 4.64], *p*=0.006), anxiety (*b*=5.33, 95%CI [2.53, 8.12], *p*=0.001), and anxious distress (*b*=6.93, 95%CI [3.47, 10.40], *p*=0.001), and lower life satisfaction (*b*=-3.20, 95%CI [-5.68, −0.71], *p*=0.012) and engagement (*b*=3.48, 95%CI [1.40, 5.56], *p*=0.001). CPSS scores were not associated with loneliness (*b*=4.40, 95%CI [-0.51, 9.31], *p*=0.078), self-care (*b*=-1.59, 95%CI [-6.80, 3.62], *p*=0.547), or social relationships (*b*=-2.04, 95%CI [-4.59, −0.51], *p*=0.117). Floor effects were minimal (0.86%), with no ceiling effects.

**Conclusions:** The CPSS is a valid instrument for assessing multidimensional stress among care partners of individuals with neurocognitive disorders.

## Introduction

Dementia continues to affect a growing number of older individuals, with projections estimating nearly 1 million persons living with dementia (PLWD) in Canada by 2030 and over 1.7 million by 2050.^1^ The role of family and friend care partners is becoming ever more critical. Care partners provide an estimated 470 million hours of support each year, an effort valued in the billions of dollars when translated into wage equivalents.^1^ This immense and growing care partner burden, which is expected to nearly triple over the next 30 years,^1^ underscores the need for comprehensive tools to assess the multifaceted challenges experienced by these caregivers.

The care partner experience is inherently multidimensional, encompassing not only the cognitive and behavioural challenges presented by PLWD, but also substantial personal, social, and financial effects arising from the cumulative demands of long-term caregiving. Current tools do not capture the full breadth of stress experienced by family and friend care partners. For example, most existing instruments assess isolated dimensions of stress, such as emotional or physical strain, with limited capacity to reflect the broader and evolving stress profile experienced by care partners.^2,3^ While some tools, such as the Caregiver Burden Inventory (CBI),^4^ offer a multidimensional structure (e.g., time-dependence, developmental, physical, social, and emotional burden), the CBI was developed over three decades ago and may lack the psychometric rigour, normative data, and contemporary relevance needed for today’s caregiving contexts. More recent tools aimed at family and friend care partners have improved in certain areas, but many remain limited in scope or lack validation for use in large, community-based samples.^5–7^ Existing scales such as the Zarit Burden Interview have contributed significantly to caregiver research,^8^ but were developed in a different era and may not fully reflect the evolving needs, identities, and contexts of today’s care partners. There remains a need for a contemporary, psychometrically sound instrument that frames caregiving challenges in terms of stress rather than burden, accommodates diverse care relationships and cultural backgrounds, and is optimized for digital administration in large-scale, community-based research.^9^

Recognizing these gaps, we developed a new Care Partner Stress Scale (CPSS) tailored specifically to assess the multidimensional realities of stress experienced by family and friend care partners of older adults with neurocognitive disorders and neurodegenerative disease (NCD/ND). The scale is designed to capture a broad spectrum of stressors including those stemming from the cognitive, functional, and behavioural changes experienced by individuals living with NCD/ND as well as psychological symptoms, practical and personal challenges, financial hardships, legal concerns, and cultural or gender role expectations faced by family and friend care partners. Here, we aim to validate this instrument by examining internal consistency and construct validity in a Canada-wide sample.

## Methods

### Study design

The Canadian Platform for Research Online to Investigate Health, Quality of Life, Cognition, Behaviour, Function, and Caregiving in Aging (CAN-PROTECT) study is a digital epidemiology platform launched in March 2023^10^ to explore risk and resilience in brain aging among community dwelling adults.^11–16^ This longitudinal online observational cohort study collects data annually on demographics, cognitive and neuropsychological test scores, behaviour and past mental health history, function, physical health, lifestyle, and quality of life (QoL) amongst others. Consistent with the goal of collecting data across the lifespan, all adults aged 18 years and older are eligible for enrolment. Participants with an established diagnosis of dementia were excluded.

In addition to the core assessments, CAN-PROTECT incorporates a dedicated caregiver sub-study. This caregiver component is designed to gain insights into the health and experiences of individuals providing care to persons with mild cognitive impairment (MCI) or dementia. Participants who report being current or past caregivers can enrol in the caregiver sub-study, which includes both care partners (e.g., family members, friends) and paid caregiving professionals (e.g., home care staff, health care or personal care aids, recreational and occupational therapists, physicians, nurses, etc.). In addition to completing CAN-PROTECT testing and assessments for the caregiver substudy, family and friend care partners complete two additional questionnaires examining the experience of stress in their caregiving role (i.e., the CPSS) and identifying available supports and resources. All caregivers are also CAN-PROTECT participants, providing comprehensive longitudinal caregiver evaluations over the same 20-year timeline as the main study.

CAN-PROTECT adheres to the ethical principles outlined in the Tri-Council Policy Statement: Ethical Conduct for Research Involving Humans (TCPS 2); the Health Canada Food and Drug Regulations (Division 5, Part C); the International Council for Harmonization of Technical Requirements for Pharmaceuticals for Human Use (ICH) E6 guideline for Good Clinical Practice; and the provisions and regulations of the Alberta Health Information Act (RSA 2000, c H-5). The study protocol was approved by the Conjoint Health Research Ethics Board (CHREB) (Ethics ID# REB21-1065) at the University of Calgary, Calgary, Alberta, Canada. All participants provided informed consent electronically for participation and publication. Participant privacy was protected with coded research identifiers, and no personally identifying information was linked to the data used in this analysis. Data were collected from March 2023 to February 2025.

### CPSS development

#### Domain and item selection

Developed specifically for CAN-PROTECT, the CPSS was informed by clinical experience working with care partners and the various efforts at identifying and addressing stress in this group. First, relevant domains contributing to stress were identified and informally vetted by a group of care partners. Then, questions were developed to populate each category. The cognitive domain included those affected in dementia (memory, language, attention, visuospatial function, executive function). The behavioural domain included the major neuropsychiatric symptom domains affected in dementia (apathy, affective symptoms, impulse dyscontrol, social inappropriateness, psychosis). Efforts were made to harmonize behavioural questions with validated diagnostic criteria for behavioural syndromes in neurocognitive disorders, i.e., apathy,^17^ agitation,^18^ psychosis,^19, 20^ and mild behavioural impairment (MBI).^21^ The functional domain included examples of instrumental and basic activities of daily living, and tasks care partners often assist with (e.g., attending appointments). Unmet Needs and Emotional Impact was populated with questions identifying care partner feelings and experiences that were insufficiently attended to because of caregiving duties (e.g., self-care or personal time). Work Interference and Financial Strain included questions about missing work and concerns over the costs of care. Family Interference and Interpersonal Conflict included questions about the impact of caregiving on family and personal relationships. Situational Perception domain questions were developed to identify stress associated with an external locus of control as a care partner as well as gender and cultural expectations around caregiving. Care partners reviewed all questions to ensure face validity, over two iterations of verbal and/or electronic feedback. The final scale included 46 items across the seven identified domains.

#### Structure and scoring

A six-point Likert scale was implemented to measure the frequency of stressors experienced by family and friend care partners over the past month for each item. Responses range from 0 (“Never”) to 6 (“Constantly”), with higher scores indicating greater frequency of stressful experiences. The total CPSS score is obtained by summing all 46 items, with possible scores ranging from 0 to 276, providing an overall measure of care partner stress.

The scale encompasses a broad range of care partner stress-related questions with a different number of questions in each domain: 1) Cognitive Decline in the care recipient (5 items, e.g., forgetfulness, inattention, difficulties with decision making, score range 0-30); 2) Behavioural Changes in the care recipient (8 items, e.g., apathy, verbal and physical aggression, psychosis, score range 0-48); 3) Functional Impairment in the care recipient (7 items, e.g., assisting with activities of daily living, managing caregiving tasks, score range 0-42); 4) Unmet Needs and Emotional Impact (6 items, e.g., difficulty maintaining self-care, limited social life, score range 0-36); 5) Work Interference and Financial Strain (4 items, e.g., missed work, financial hardship, score range 0-24); 6) Family Interference and Interpersonal Conflict (5 items, e.g., disagreements over care decisions, lack of time for one’s own family, score range 0-30); and 7) Situational Perception (11 items, e.g., inadequate support from family or formal services, gender or cultural expectations of caregiving, score range 0-66) (Figure 1; Supplementary Figure 1).

**Figure 1.**
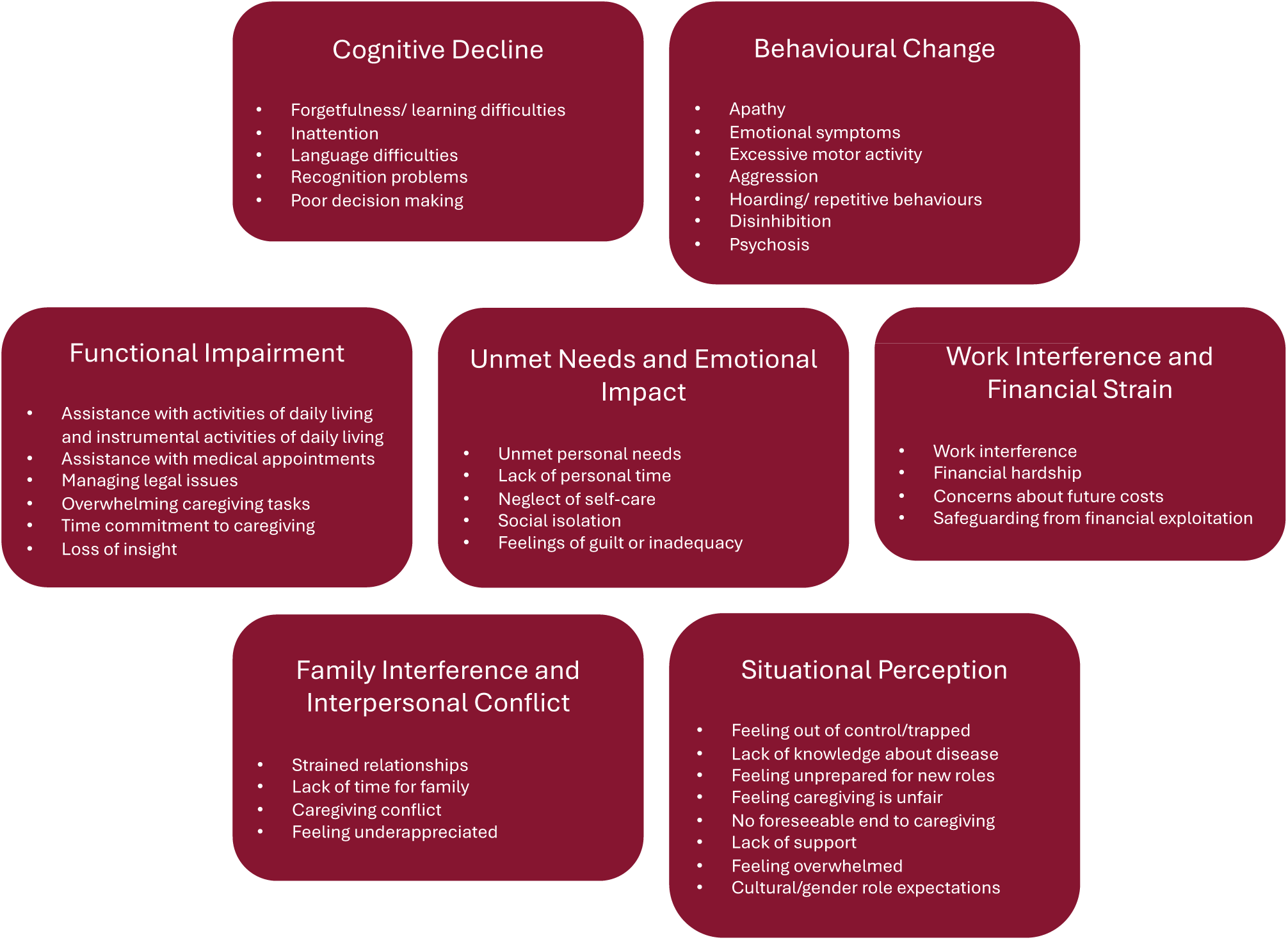
Conceptual overview of Care Partner Stress Scale domains with paraphrased item content. The Care Partner Stress Scale assesses care partner-related stress across seven domains. This figure summarizes each domain and includes paraphrased descriptors of the types of stressors assessed. The full instrument can be obtained from the authors under appropriate terms.

### Participant selection

Study flow for data analysis is shown in Figure 2. Care partner participants enrolled between March 2023 and February 2025 who had completed the CPSS were considered for this validation study. Participants were included if they had completed the Patient Health Questionnaire (PHQ-9),^22^ Generalized Anxiety Disorder-7 (GAD-7) scale,^23^ Calgary Anxious Distress Inventory (CADI),^10^ Quality of Life and Function Five Domain Scale (QFS-5; Self-care, Social Relationships, and Life Satisfaction domains),^12^ University of California Los Angeles Loneliness Scale 3 (UCLA-LS-3),^24^ and Inventory of Depressive Symptomatology Self Report Life Engagement Subscale 10 (IDS-SR 10).^25^

**Figure 2.**
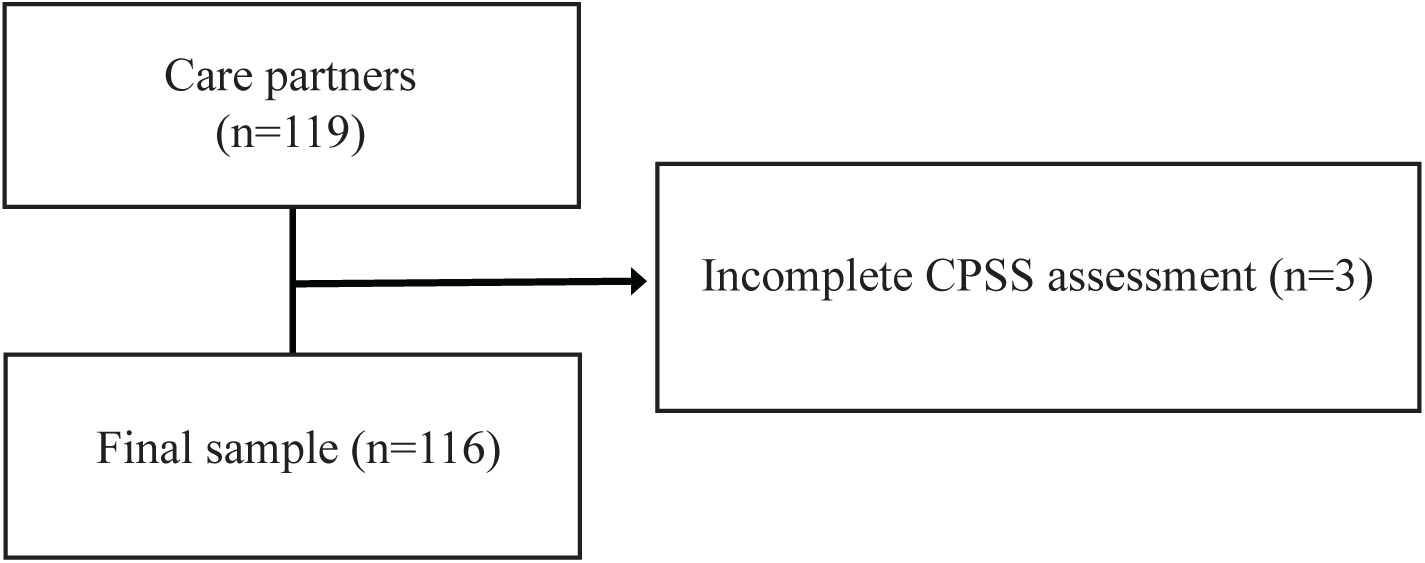
Flowchart of CAN-PROTECT care partner participants included for analysis. CAN-PROTECT: Canadian Platform for Research Online to Investigate Health, Quality of Life, Cognition, Behaviour, Function, and Caregiving in Aging; CPSS: Care Partner Stress Scale.

### Statistical analyses

Demographic characteristics, including age, sex, origin, and years of education, were of interest for this study. Handedness was also included as a variable expected to be unrelated to care partner stress, supporting later analysis of discriminant validity. Clinical data included scores on the CPSS, PHQ-9, CADI, GAD-7, QFS-5 (Self-care, Social Relationships, and Life Satisfaction domains), UCLA-LS-3, and IDS-SR 10. Continuous variables were summarized using means, standard deviation (SD), and ranges, while categorical variables were reported as counts and percentages. Since the number of items differed across CPSS domains, domain scores were calculated as the average of the respective item scores, ensuring comparability across domains. All participants were included in these calculations. Endorsement rates were calculated at the item and domain levels as the proportion of participants reporting a score greater than zero. In addition to overall means (calculated across all responses, including zeros), endorsed item means were calculated at both the item and domain levels using only items scored greater than zero. This approach provides an estimate of average severity conditional on endorsement, allowing us to describe stress levels specifically among those for whom each stressor was relevant.

Item-level analysis was conducted to evaluate the contribution of individual items to the overall scale. Internal consistency was assessed using Cronbach’s alpha, with higher coefficients indicating greater reliability and alignment of items with the intended construct of care partner stress. Item-total correlations were calculated to assess the relationship between each individual item and the total score of the questionnaire. Adjusted correlations were then examined to identify potentially irrelevant items that might undermine scale reliability. Items with higher adjusted correlations were considered more strongly aligned with the construct of care partner stress. Additionally, Guttman’s G6 coefficient was calculated to provide an alternative estimate of internal consistency and latent construct strength.^26^ Guttman’s G6 coefficient provides a less biased reliability estimate in cases where the assumptions of unidimensionality required for Cronbach’s alpha may not be fully met, offering further insight into the degree to which the items reflect a common latent construct.

Test-retest reliability and inter-rater reliability were not evaluated in this study. Assessments, including the CPSS, are administered annually in CAN-PROTECT. The lengthy interval between assessments precluded effective test-retest evaluation, which typically requires shorter intervals to assess instrument stability. Hence, other study designs are required to establish test-retest reliability. Furthermore, since the CPSS is a self-reported measure intended to measure caregiver self-perception of stress, inter-rater reliability was not evaluated.

Criterion validity could not be assessed because no established alternative measure of care partner stress was available for comparison. More broadly, there are few widely used and validated instruments specifically designed to assess stress across multiple domains (rather than burden) in care partners of adults with NCD/ND. However, we examined convergent and discriminant validity for the CPSS to establish construct validity. Convergent validity was assessed by examining the linear relationship between the CPSS total score and other measures known to be associated with care partner stress, including PHQ-9, GAD-7, CADI, QFS-5 (Self-Care, Social Relationships, and Life Satisfaction domains), UCLA-LS-3, and IDS-SR 10 total scores. These instruments measure depression, anxiety, anxious-distress, QoL, loneliness, and life engagement, each considered relevant to care partner stress. Discriminant validity was evaluated by examining the relationship between the CPSS total score and handedness (right, left), as we did not expect CPSS to be correlated with handedness. Ambidextrous individuals were pooled with the left-handed group for analysis, and right-handed participants were set as the reference group. All models were estimated using linear regressions with CPSS total score as the outcome variable and no covariates. To minimize the influence of extreme values on regression estimates, CPSS total scores were Winsorized at the 5th and 95th percentiles for construct validity analyses.

The PHQ-9 is a widely used 9-item measure of depressive symptoms, assessing frequency over the past two weeks using a 4-point Likert scale ranging from 0 (“Not at all”) to 3 (“Nearly every day”).^22^ Higher total scores indicate greater symptom severity. Similarly, the GAD-7 is a 7-item measure assessing frequency of symptoms of generalized anxiety disorder over the past two weeks, also using a 4-point Likert scale from 0 (“Not at all”) to 3 (“Nearly every day”).^23^ Higher scores on the GAD-7 reflect greater severity of anxiety symptoms. The CADI is a 5-item measure assessing symptoms of anxious distress over the past two weeks. Items assess the severity of specific symptoms (e.g., feeling keyed up or tense, sense of foreboding) and are rated on a 4-point Likert scale from 0 (“Not present”) to 3 (“Severe - very marked or prominent, or dramatic”). Higher total scores indicate greater severity of anxious distress. The QFS-5 is a newly developed 25-item measure evaluating QoL and functional abilities across five domains (Productivity, Self-care, Social Relationships, Life Satisfaction, and Leisure and Activities).^12^ Each item is rated based on abilities using a 4-point Likert scale from 0 (“Never”) to 3 (“Always”), with higher scores indicating better QoL. Of interest to this study were the QSF-5 Self-care, Social Relationships, and Life Satisfaction domains. Unlike other QoL measures, the QFS-5 excludes specific health-related items (e.g., depression, pain) to avoid potential rating confounds in individuals with mental or physical health conditions.^12^ The UCLA-LS-3 is a brief, validated measure assessing feelings of loneliness and social isolation.^24^ The scale includes three items rated on a 3-point Likert scale from 0 (“Hardly ever”) to 2 (“Often”), with higher scores indicating greater loneliness. This shortened version maintains strong psychometric properties, demonstrating good internal consistency and convergent validity with longer loneliness measures, and is well-suited for large-scale studies.^27, 28^ The IDS-SR 10 is a 10-item measure, derived from the original IDS-SR, assessing life engagement over the past seven days.^25^ Developed in accordance with the 4-domain model of life engagement,^29^ the scale assesses emotional (affect/mood), physical (energy/motivation), social (involvement/interest), and cognitive (alertness/thinking) domains.^25^ Each item is rated on a 4-point Likert scale from 0 to 3, with higher scores indicating lower life engagement.

Content validity and data completeness were evaluated through examination of floor and ceiling effects. Following established guidelines, floor or ceiling effects were considered present if more than 15% of participants achieved the lowest or highest possible score, respectively.^30, 31^

## Results

### Participant demographics and characteristics

Participant characteristics are summarized in Table 1. The CAN-PROTECT care partner sample consists of older adults who are predominantly female (87.1%), highly educated (mean±SD years of education=16.75±4.35), and of North American and/or European origins. Care partners reported moderate levels of stress (107.02±47.15, range: 0-226), with the highest levels observed in the Cognitive Decline domain, reflecting the significant burden of cognitive impairment in the care recipient and the associated challenges of managing progressive memory loss and impaired decision-making. Other notable sources of stress by domain included Functional Impairment of the care recipient, as well as Unmet Needs and Emotional Impact, and Situational Perception of the care partner. The top items contributing to care partner stress were related to the care recipient’s forgetfulness or difficulties with learning, the amount of time spent on caregiving obligations, managing instrumental activities of daily living, feeling unable to control the situation, the care recipient’s difficulty with decision-making, and attending or assisting with medical appointments (Figure 3).

**Figure 3.**
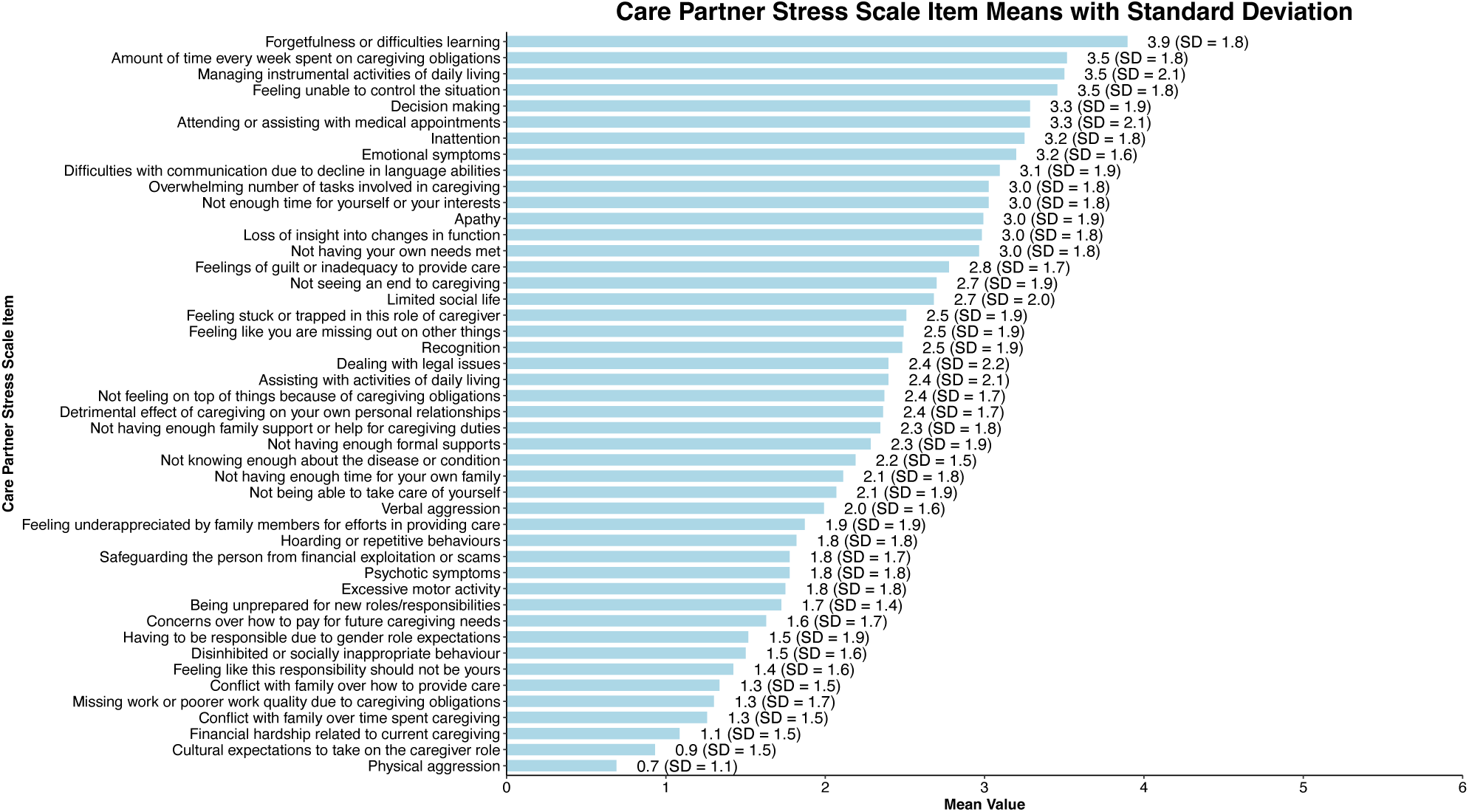
Mean scores and standard deviations for individual items of the Care Partner Stress Scale. Mean scores (± standard deviation) for each Care Partner Stress Scale item are displayed. Higher mean values indicate more frequent stress related to the specified caregiving challenge over the past month. Items are ordered from highest to lowest mean score. *SD*: Standard deviation.

**Table 1.**
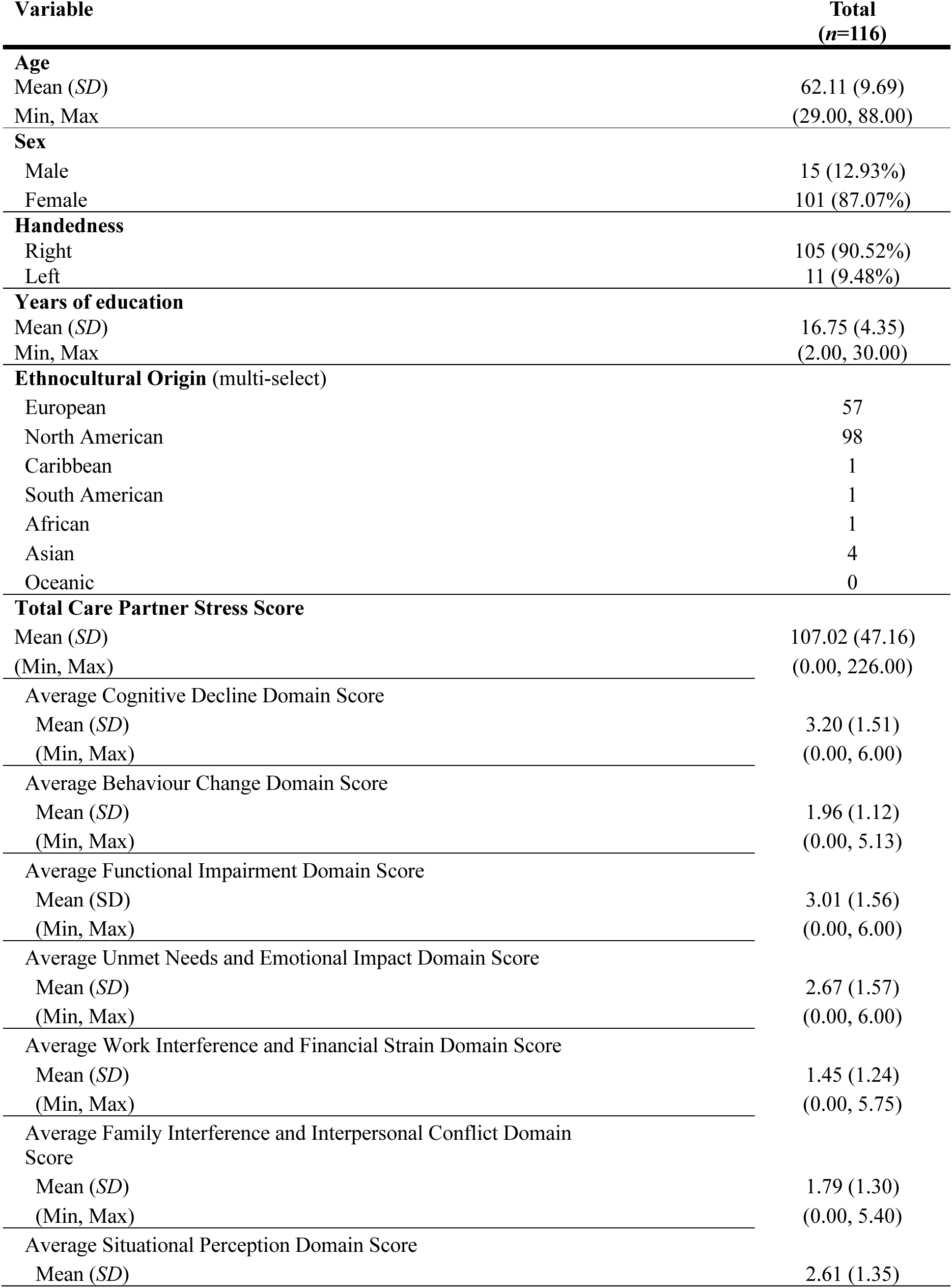
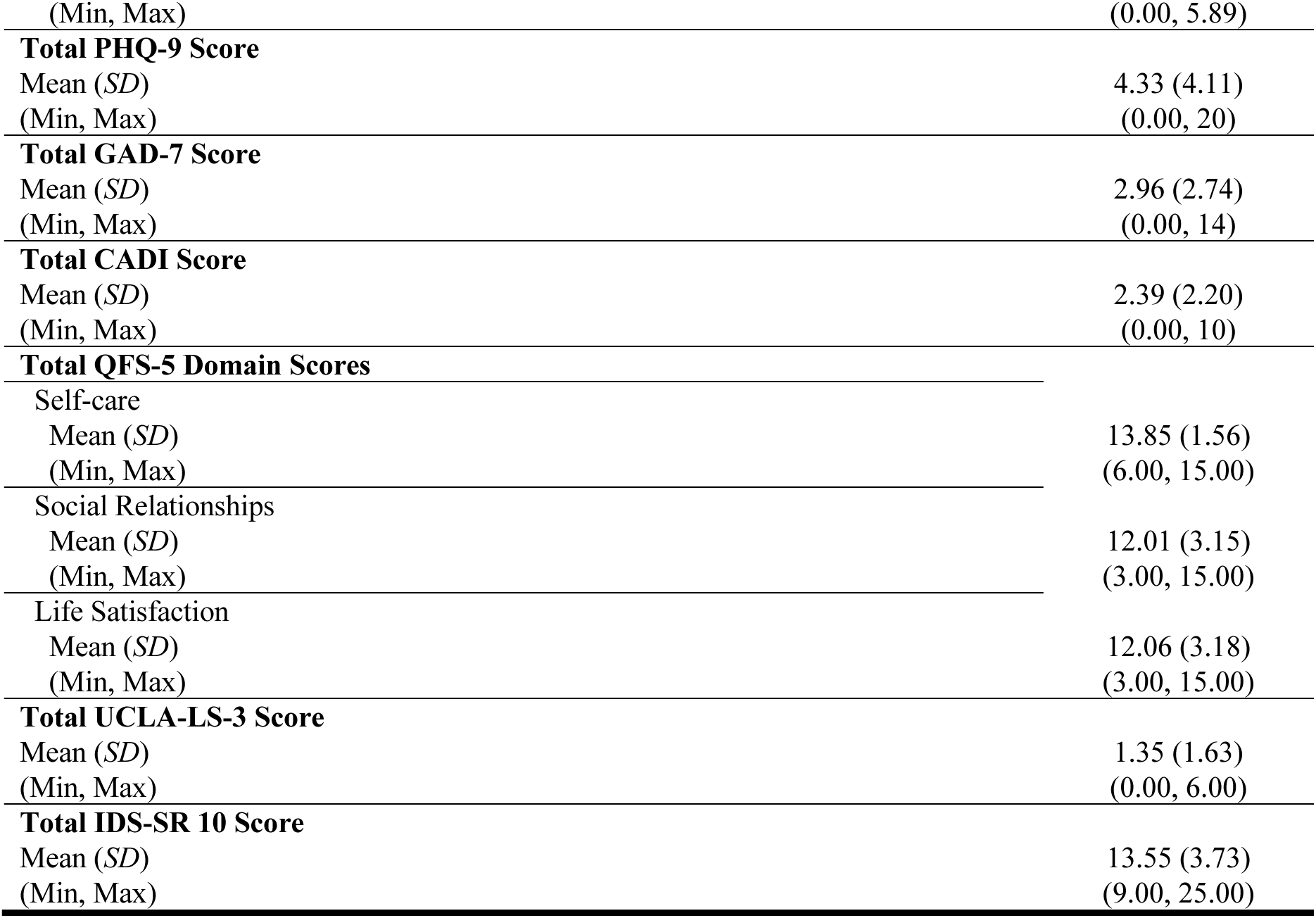
Participant Characteristics. PHQ-9: Patient Health Questionnaire, GAD-7: Generalized Anxiety Disorder-7, CADI: Calgary Anxious-Distress Inventory, QFS-5: Quality of Life and Function Five Domain Self Report, UCLA-LS-3: University of California Los Angeles Loneliness Scale 3, IDS-SR 10: Inventory of Depressive Symptomatology-Self Report 10; SD: Standard deviation.

To further examine care partner stress, we distinguished between overall mean scores (calculated across all responses, including zeros) and endorsed item means, calculated at both the item and domain levels using only items scored greater than zero. Endorsement rates were high across domains, with 89.66% of participants endorsing stress related to the Cognitive Decline domain, 69.81% related to Behavioural Change, 84.73% related to Functional Impairment, 84.78% related to Unmet Needs and Emotional Impact, 56.90% related to Work Interference and Financial Strain, 69.92% related to Family Interference and Interpersonal Conflict, and 74.31% related to Situational Perception. When examining mean scores of endorsed items, the Cognitive Decline domain remained the highest source of stress (3.56±1.62), followed by Functional Impairment (3.55±1.70), Unmet Needs and Emotional Impact (3.14±1.59), Situational Perception (2.83±1.51), Behavioural Change (2.71±1.45), Family Interference and Interpersonal Conflict (2.55±1.46), and Work Interference and Financial Strain (2.54±1.47). A similar pattern was observed at the item level. When considering only non-zero individual items, the top contributors to care partner stress were care recipient forgetfulness or difficulties with learning, managing instrumental activities of daily living, the amount of time spent on caregiving obligations, feeling unable to control the situation, and attending or assisting with medical appointments (Figure 4).

**Figure 4.**
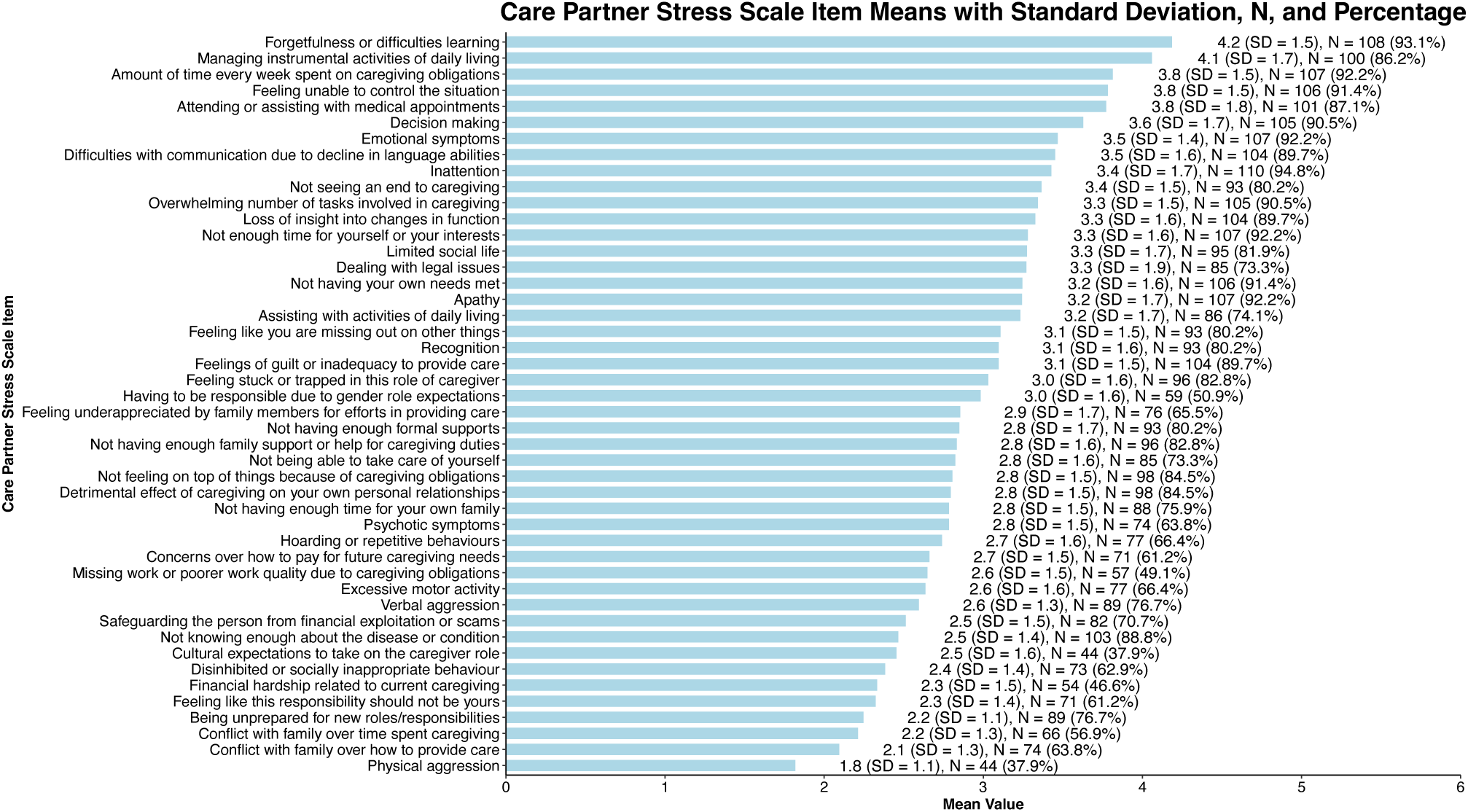
Mean scores of Care Partner Stress Scale items among items endorsed. Mean scores (± standard deviation) are shown for each Care Partner Stress Scale item endorsed (i.e., scored > zero). The number and percentage of participants endorsing each item are also reported. Higher mean values reflect more frequent stress related to the caregiving challenge. *SD*: standard deviation, *N*: number of endorsers.

Care partners in this sample reported relatively low levels of depressive symptoms, anxiety, loneliness, and life engagement while overall QoL was moderate to high.

### CPSS item descriptive statistics and reliability

Means, SDs, and item-total correlations (unadjusted and adjusted) for each item are presented in Table 2. Item-total correlations reflect the relationship between each item score and the total score of all other items excluding the item itself. All items demonstrated acceptable adjusted item-total correlations above the commonly used threshold of 0.30,^32^ with the lowest correlation being 0.31. This suggests that no items needed to be deleted or significantly modified, as all contributed to the overall construct of care partner stress. Most items exhibited moderate to strong correlations (>0.50),^33^ indicating good internal consistency. A few items had low frequency of endorsement in this sample (e.g., physical aggression, psychotic symptoms, cultural expectations to take on the caregiver role) and thus, were closer to the lower bound of acceptability. However, these items still met the minimum threshold, suggesting they do not substantially detract from the scale’s reliability.

**Table 2.** Descriptive statistics and item-total correlations for the Care Partner Stress Scale. Means, *SD*, and *r* are presented for each item. Unadjusted *r* values represent the correlation between each item and the total Care Partner Stress Scale score, while adjusted *r* values reflect the correlation between each item and the total score excluding the item itself (corrected item- total correlation). Higher adjusted correlations indicate stronger alignment of the item with the overall construct of care partner stress. *SD*: Standard deviation; *r*: Pearson correlation coefficient.

| Item | Mean | *SD* | *r*<br>(unadjusted) | *r*<br>(adjusted) |
| --- | --- | --- | --- | --- |
| Forgetfulness or difficulties learning | 3.90 | 1.8 | 0.57 | 0.56 |
| Inattention | 3.25 | 1.8 | 0.63 | 0.62 |
| Difficulties with communication due to decline in language abilities | 3.09 | 1.9 | 0.51 | 0.50 |
| Recognition | 2.48 | 1.9 | 0.61 | 0.61 |
| Decision making | 3.28 | 1.9 | 0.60 | 0.59 |
| Apathy | 2.99 | 1.9 | 0.65 | 0.64 |
| Emotional symptoms | 3.20 | 1.6 | 0.58 | 0.58 |
| Excessive motor activity | 1.75 | 1.8 | 0.55 | 0.55 |
| Verbal aggression | 1.99 | 1.6 | 0.36 | 0.36 |
| Physical aggression | 0.69 | 1.1 | 0.32 | 0.32 |
| Hoarding or repetitive behaviours | 1.82 | 1.8 | 0.54 | 0.53 |
| Disinhibited or socially inappropriate behaviour | 1.50 | 1.6 | 0.39 | 0.38 |
| Psychotic symptoms | 1.78 | 1.8 | 0.32 | 0.31 |
| Assisting with activities of daily living | 2.40 | 2.1 | 0.66 | 0.65 |
| Managing instrumental activities of daily living | 3.50 | 2.1 | 0.66 | 0.64 |
| Attending or assisting with medical appointments | 3.28 | 2.1 | 0.64 | 0.62 |
| Dealing with legal issues | 2.40 | 2.2 | 0.47 | 0.45 |
| Overwhelming number of tasks involved in caregiving | 3.03 | 1.8 | 0.72 | 0.71 |
| Amount of time every week spent on caregiving obligations | 3.52 | 1.8 | 0.73 | 0.72 |
| Loss of insight into changes in function | 2.98 | 1.8 | 0.64 | 0.63 |
| Not having your own needs met | 2.97 | 1.8 | 0.70 | 0.70 |
| Not enough time for yourself or your interests | 3.03 | 1.8 | 0.74 | 0.74 |
| Not being able to take care of yourself | 2.07 | 1.9 | 0.73 | 0.73 |
| Limited social life | 2.68 | 2.0 | 0.67 | 0.67 |
| Feeling like you are missing out on other things | 2.49 | 1.9 | 0.68 | 0.68 |
| Feelings of guilt or inadequacy to provide care | 2.78 | 1.7 | 0.71 | 0.71 |
| Missing work or poorer work quality due to caregiving obligations | 1.30 | 1.7 | 0.55 | 0.55 |
| Financial hardship related to current caregiving | 1.09 | 1.5 | 0.54 | 0.55 |
| Concerns over how to pay for future caregiving needs | 1.63 | 1.7 | 0.52 | 0.52 |
| Safeguarding the person from financial exploitation or scams | 1.78 | 1.7 | 0.38 | 0.36 |
| Detrimental effect of caregiving on your own personal relationships | 2.36 | 1.7 | 0.74 | 0.75 |
| Not having enough time for your own family | 2.11 | 1.8 | 0.72 | 0.73 |
| Conflict with family over how to provide care | 1.34 | 1.5 | 0.43 | 0.43 |
| Conflict with family over time spent caregiving | 1.26 | 1.5 | 0.55 | 0.56 |
| Feeling underappreciated by family members for efforts in providing care | 1.87 | 1.9 | 0.54 | 0.54 |
| Feeling unable to control the situation | 3.46 | 1.8 | 0.72 | 0.71 |
| Not knowing enough about the disease or condition | 2.19 | 1.5 | 0.40 | 0.40 |
| Being unprepared for new roles/responsibilities | 1.72 | 1.4 | 0.43 | 0.43 |
| Feeling like this responsibility should not be yours | 1.42 | 1.6 | 0.48 | 0.48 |
| Feeling stuck or trapped in this role of caregiver | 2.51 | 1.9 | 0.67 | 0.67 |
| Not seeing an end to caregiving | 2.70 | 1.9 | 0.64 | 0.64 |
| Not having enough family support or help for caregiving duties | 2.34 | 1.8 | 0.62 | 0.62 |
| Not having enough formal supports (e.g., home care, day program, or respite care) | 2.28 | 1.9 | 0.54 | 0.53 |
| Not feeling on top of things because of caregiving obligations | 2.37 | 1.7 | 0.74 | 0.75 |
| Having to be responsible due to gender role expectations | 1.52 | 1.9 | 0.41 | 0.40 |
| Cultural expectations to take on the caregiver role | 0.93 | 1.5 | 0.33 | 0.32 |

The CPSS demonstrated excellent internal consistency, with a Cronbach’s alpha of 0.96 (95% CI: 0.94-0.97), exceeding the commonly accepted threshold of 0.70 for good reliability.^34^ The standardized alpha was also high (0.95), further supporting reliability. The average inter-item correlation was 0.31, aligning with recommended values (0.15-0.50)^35^ for well-constructed psychological scales. Additionally, the Guttman’s G6 coefficient was 0.98, indicating strong latent construct consistency. Item deletion analyses showed that removing any individual item did not meaningfully change the overall reliability (all α = 0.95-0.96), further supporting the robustness of the CPSS. These findings indicate that the scale reliably measures care partner stress as a unified construct and that all items contribute meaningfully to the assessment.

### Convergent and discriminant validity

To assess convergent validity, we examined correlations between the CPSS and established measures of psychological distress and well-being (Figure 5). Higher care partner stress scores were significantly associated with greater symptoms of depression (PHQ-9: *b* = 2.73, 95%CI [0.82, 4.64], *p* = 0.006), anxiety (GAD-7: *b* = 5.33, 95%CI [2.53, 8.12], *p* < 0.001), and anxious distress (CADI: *b* = 6.93, 95%CI [3.47, 10.40], *p* < 0.001), as well as lower life satisfaction (QSF-5 Life Satisfaction domain: *b* = −3.20, 95%CI [−5.68, −0.71], *p* = 0.012) and life engagement (IDS-SR 10: *b* = 3.48, 95%CI [1.40, 5.56], *p* = 0.001). Associations with loneliness (UCLA-LS-3: *b* = 4.40, 95%CI [−0.51, 9.31], *p* = 0.078), self-care (QFS-5 Self-care domain: *b* = −1.59, 95%CI [−6.80, 3.62], *p* = 0.547), and social relationships (QFS-5 Social Relationships domain: *b* = −2.04, 95%CI [−4.59, 0.51], *p* = 0.117) were in the expected direction but did not reach statistical significance. To assess discriminant validity, we examined the relationship between the CPSS and handedness. Regression analysis revealed that CPSS scores were not significantly associated with handedness (*b* = −1.54, 95%CI [−29.15, 26.08], *p* = 0.912).

**Figure 5.**
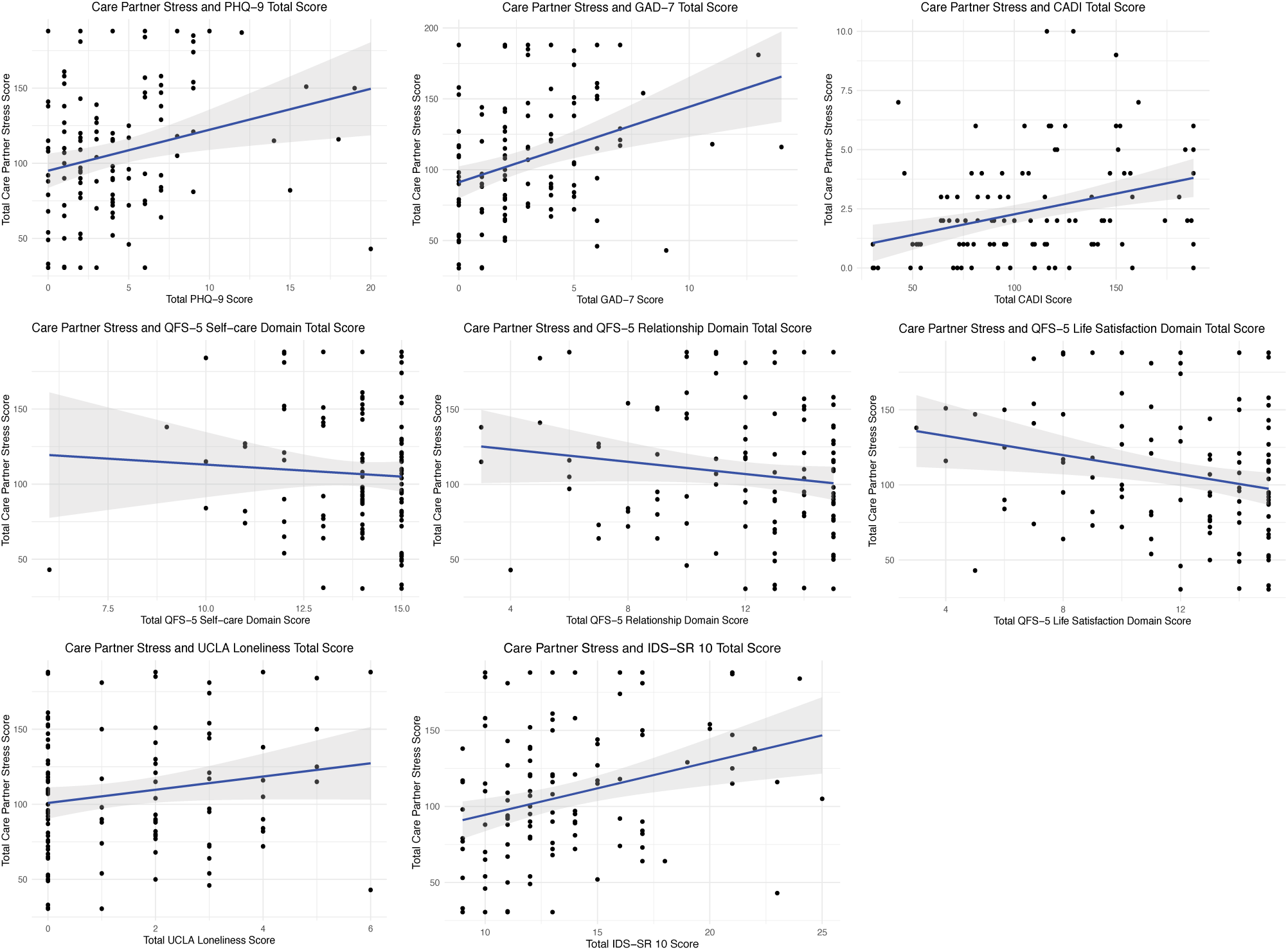
Associations between Care Partner Stress Scale scores and psychological distress and well-being measures. Scatterplots with fitted regression lines (±95% confidence intervals) display the relationship between Care Partner Stress Scale scores and measures of depression, anxiety, anxious distress, loneliness, quality of life, and life engagement. Higher Care Partner Stress Scale scores indicate greater perceived caregiving stress. PHQ-9: Patient Health Questionnaire, GAD-7: Generalized Anxiety Disorder-7, CADI: Calgary Anxious-Distress Inventory, QFS-5: Quality of Life and Function Five Domain Self Report, UCLA-LS-3: University of California Los Angeles Loneliness Scale 3, IDS-SR 10: Inventory of Depressive Symptomatology-Self Report 10.

Our analysis revealed a minimal floor effect (0.86%), with only one participant scoring at the lowest possible value. In contrast, no ceiling effects (0%) were observed, as no participants scored at the highest possible value. This suggests that the CPSS adequately captures variations in stress levels without substantial floor or ceiling limitations.

## Discussion

This study provides initial validation evidence for the CPSS, a novel instrument designed to assess the multidimensional nature of stress among care partners of individuals living with NCD/ND. The CPSS demonstrated excellent internal consistency and moderate to strong item-total correlations, suggesting that all items contributed meaningfully to the overarching construct of care partner stress. Item- and domain-level endorsements confirmed that stress related to the care recipient’s cognitive decline and functional impairment were particularly prominent, aligning with previous findings that increasing dependence and functional decline are major predictors of care partner burden.^36^ Additionally, stress due to unmet needs and the emotional impact of caregiving were highly endorsed, echoing international survey data showing that a majority of care partners report significant unmet needs in managing stress.^37^ These findings support the CPSS as a psychometrically robust instrument capable of capturing the complex and varied challenges experienced by care partners of persons living with neurocognitive disorders.

Importantly, the specific domains represented in the CPSS are consistent with known sources of caregiver stress. For instance, care partners frequently face unpredictable neuropsychiatric symptoms and disruptive behaviours in the care recipient, identified as some of the most distressing aspects of dementia caregiving, alongside the emotional toll of juggling multiple caregiving roles and navigating family conflict.^38, 39^

Convergent validity was supported by significant associations between CPSS scores and measures of depression, generalized anxiety, anxious distress, life satisfaction, and life engagement. This pattern aligns with substantial evidence linking greater caregiver stress to poorer mental health and well-being.^40–43^ Discriminant validity was demonstrated through a lack of association between CPSS score and handedness, indicating that the scale is not merely capturing generalized negative responding. CPSS score distributions also showed minimal floor effects and no ceiling effects, indicating strong sensitivity across the stress continuum, and suggesting the scale can detect both low and high levels of stress without clustering at the extremes. This is a notable advantage over some shorter burden tools, which can exhibit restricted range or skewed distributions in certain populations. By encompassing multiple domains of care partner stress, the CPSS likely mitigates such problems and can detect both subtle and pronounced changes in stress levels over time.

This validation effort is timely and important for several reasons. First, it provides researchers and clinicians with a contemporary and robust tool that extends beyond traditional assessments of care partner burden. Existing burden scales tend to emphasize emotional, physical, or financial strain, but may not fully reflect the nuanced and evolving stressors faced by today’s care partners. In contrast, the multi-domain design of the CPSS captures a wider array of care partner challenges, including practical, emotional, and gender and sociocultural factors that have been under-recognized in earlier tools. This approach is critical given emerging evidence that care partner burden is not a uniform experience, but one shaped by a complex interplay of individual, relational, and contextual factors including caregiver characteristics, care recipient needs, and the sociocultural context in which caregiving occurs.^44^ Second, given the projected rise in dementia prevalence and the corresponding increase in the number of care partners, a comprehensive understanding of care partner stress is essential for informing targeted interventions and policy measures. Recognizing caregiving as a shared societal responsibility rather than an individual burden demands instruments that reflect the full scope of care partner experiences.^1^ The CPSS can contribute meaningfully to this shift by identifying specific stress domains where targeted support is most needed. Indeed, policy frameworks such as the World Health Organization Global Action Plan on the Public Health Response to Dementia, emphasize care partner support as an essential pillar of dementia care systems.^45^

A key strength of the CPSS lies in the caregiver-informed development process, which involved collaboration with family and friend care partners to ensure relevance and face validity. The CPSS incorporates items such as work interference, situational perception, and cultural or gender-based expectations, reflecting the unique realities of unpaid care partners. Notably, care partners of dependent older adults experience stress differently from paid caregivers,^46^ potentially due to contextual and relational factors specific to family and friend caregiver experiences. By directly involving care partners in the development and vetting of items, the CPSS ensures that each domain resonates with real-world caregiving experiences in care partner contexts. This results in a scale that is sensitive, contextually grounded, and capable of identifying specific sources of stress that may otherwise go unrecognized.

As the prevalence of dementia increases, more family members and friends will assume caregiving responsibilities. Understanding the full spectrum of stressors faced is essential for guiding the design of responsive interventions, the provision of tailored support services, and the formulation of policies that reflect the realities of family and friend caregiving. The CPSS addresses this need by providing a finely tuned tool that captures the complexity of the care partner experience and helps identify where support is most urgently needed.

Strengths of our study include the use of a large, geographically diverse sample. The use of a Canada-wide sample enhances the generalizability of the scale across different populations and both rural and urban settings. As clinical and research assessments increasingly transition to digital platforms, implementing reliable scales that participants can complete online is essential. Additionally, the rigorous psychometric evaluation, including internal consistency, item-total correlations, and associations with various measures of psychological distress and well-being, provides strong evidence of reliability and validity. Notably, the pattern of associations observed with the CPSS closely mirrors those reported in studies using established caregiver burden scales,^8^ reinforcing that the CPSS captures a relevant and meaningful construct. Together, these features position the CPSS as a practical and robust measure of care partner stress, with utility for both research and clinical practice.

### Limitations

Several limitations should be considered when interpreting these results. First, the CPSS has not yet been evaluated for test-retest reliability, leaving uncertainty regarding the temporal stability of scores over shorter time periods. This limitation reflects that assessments in CAN-PROTECT are completed annually; hence, other study designs with shorter follow-up intervals are required to establish test-retest reliability. Second, while inter-rater reliability was not applicable given the self-report nature of the scale, reliance solely on caregiver perceptions may introduce bias or subjective interpretation. At the same time, perceived stress is inherently subjective and is most appropriately assessed through self-report by the individual experiencing stress. Nonetheless, the absence of complementary objective indicators (e.g., physiological or biological stress markers) limits the ability to triangulate self-reported stress with external measures, an important consideration, particularly in the context of online research where the collection of such data may not be feasible. Third, although we demonstrated convergent and discriminant validity, criterion validity could not be assessed due to the absence of an established reference measure of care partner stress in the CAN-PROTECT dataset. In other words, despite the existence of caregiver burden scales, there is currently no gold-standard instrument that specifically measures stress as contemporaneously conceptualized by the CPSS. This lack of comparator constrained our ability to evaluate criterion validity directly. Additionally, the CPSS has not yet been evaluated for sensitivity to change over time or responsiveness to interventions. It remains to be seen whether the scale can detect meaningful reductions in stress following care partner support programs or other interventions, an important property for any outcome measure intended for clinical trials or longitudinal research. Future research should address these limitations to strengthen the evidence base for the CPSS and broaden the application of the scale in diverse caregiving populations.

Another methodological consideration relates to the use of Winsorization, which was applied to limit the influence of extreme CPSS scores and ensure adherence to statistical assumptions. While this approach preserved the full sample and enhanced analytic robustness, it may have attenuated variability at the upper end of the score distribution. As such, future research might consider alternative strategies for handling outliers or use longitudinal designs to determine whether extreme scores persist or regress over time, ensuring that the full range of caregiver stress is accurately captured.

Additionally, one of the convergent measures used to establish construct validity, the CADI, is a relatively new instrument. Although the CADI was selected for its conceptual relevance and low respondent burden in large-scale online research, published psychometric data remain limited. We acknowledge that the use of a newer measure with limited validation might introduce some uncertainty. Future studies should further evaluate the reliability and validity of the CADI in diverse caregiving populations to strengthen confidence in its use as an indicator of anxious distress.

Finally, while this sample provides valuable insight into care partner stress, participants may not be fully representative of the broader care partner population. Given the relatively favourable mental health profile observed, individuals experiencing greater psychological distress or with fewer support resources may have been less likely to enrol or remain engaged in the study, potentially limiting generalizability. It is also possible that the most burdened caregivers, who may experience the highest levels of stress, had limited time or capacity to participate in research. These factors may have contributed to lower prevalence of endorsement for certain items (e.g., physical aggression, psychosis) despite their clinical importance. Moreover, the prominence of cognitive stress items may partially reflect the historically cognocentric framing of dementia in both research and public discourse, which could shape how care partners interpret and report stressors. Future studies with broader enrollment will be able to address these issues. Despite these limitations, the initial validation of the CPSS represents an important step in developing a more nuanced understanding of caregiver stress.

## Conclusion

The CPSS is a reliable and valid tool for assessing multidimensional stress among care partners of PLWD or NCD/ND. The scale demonstrates strong psychometric properties and comprehensive coverage of caregiving challenges, making it a valuable resource for both research and clinical practice. By encompassing a broad range of stress domains, from the practical effects of caregiving on work and daily life to the emotional and relational strain that caregivers experience, the CPSS fills a notable gap in the toolkit available to assess family and friend care partner outcomes. Importantly, the CPSS offers a more nuanced approach to measuring care partner stress than existing tools, with the potential to meaningfully enhance the quality, specificity, and scope of future research in this area. Our findings position the CPSS as a robust measure that can be used to identify caregivers at risk of high stress, to tailor interventions to specific stress domains, and to evaluate the effectiveness of support programs aimed at alleviating caregiver burden.

Future studies should build on this work by evaluating the stability of CPSS scores over shorter intervals and examining responsiveness to interventions or changes in the caregiving context. As the number of care partners continues to grow in tandem with the aging population, having valid and sensitive instruments like the CPSS will be crucial for advancing our understanding of care partner well-being and for informing policies and interventions that ultimately improve the lives of care partners and care recipients alike.

## Acknowledgements

CAN-PROTECT is supported by Gordie Howe CARES and the Evans Family Fund through the Hotchkiss Brain Institute (HBI). The funders had no role in the study design; collection, analysis, or interpretation of data; writing of the manuscript; or the decision to submit for publication. The authors thank the CAN-PROTECT study team and research staff for their administrative and technical support. We also acknowledge the PROTECT-UK team at the University of Exeter for their collaboration and foundational work in the development of the CAN-PROTECT study platform.

## Author contributions

Daniella Vellone (Conceptualization; Data curation; Formal analysis; Methodology; Project administration; Software; Visualization; Writing – original draft; Writing – review & editing); Dylan X. Guan (Conceptualization; Data curation; Formal analysis; Methodology, Software, Writing – review & editing); Jasper F. E. Crockford (Writing – original draft; Writing – review and editing); Ibadat Warring (Writing – original draft, Writing – review and editing); Clive Ballard (Software, Writing – review & editing); Byron Creese (Software, Writing – review & editing); Anne Corbett (Software, Writing – review & editing); Ellie Pickering (Software, Writing – review & editing); Pamela Roach (Investigation, Writing – review & editing); Eric E. Smith (Investigation, Writing – review & editing); Zahinoor Ismail (Conceptualization; Funding acquisition; Methodology; Project administration; Resources; Supervision, Writing – original draft; Writing – review & editing).

## Statements and declarations

### Ethical considerations

CAN-PROTECT adheres to the Tri-Council Policy Statement: Ethical Conduct for Research Involving Humans (TCPS 2); Health Canada’s Food and Drug Regulations (Division 5, Part C); the International Council for Harmonization of Technical Requirements for Pharmaceuticals for Human Use (ICH) E6 guideline for Good Clinical Practice; and the provisions and regulations of Alberta’s Health Information Act (RSA 2000, c H-5). Study protocols were reviewed and approved by the Conjoint Health Research Ethics Board (CHREB) (Ethics ID# REB21-1065) at the University of Calgary, Calgary, Alberta, Canada. All participants provided written informed consent electronically for participation and publication. Participant privacy and data confidentiality were protected through the use of coded research identifiers, and no personally identifying information was linked to the data used in this analysis.

### Consent to participate

All CAN-PROTECT participants provided written informed consent electronically prior to the initiation of any study procedures, including consent for data collection, storage, and future research use. Informed consent included permission to collect and store longitudinal clinical assessments for research purposes.

### Consent for publication

Participants provided written informed consent authorizing the use of de-identified data for research dissemination, including scientific publication. Consent forms explicitly state that only coded, non-identifiable data will be shared and used in publications. No personally identifying information or images are included in this manuscript.

### Data availability

At this time, the authors do not have ethical or legal permission to share the study data, including de-identified data. The study is still in its early stages, and a data access committee has not yet been established, nor has a data sharing policy been finalized. Upon study completion, the authors will seek legal review and submit an amendment to the ethics board. Addressing data sharing is a priority, contingent on obtaining stable funding for all study activities. While the authors are unable to publicly post the data at this time, they are open to sharing data with qualified investigators for validation purposes. Interested researchers may contact to request access to the data. R scripts used for data cleaning and analysis in this study are available upon reasonable request to the corresponding author.

## References

1. Alzheimer Society Canada. Navigating the Path Forward for Dementia in Canada: The Landmark Study/Path/2022. 2022.

2. Zarit SH, Reever KE and Bach-Peterson J. Relatives of the impaired elderly: correlates of feelings of burden. Gerontologist 1980; 20: 649–655. DOI: 10.1093/geront/20.6.649.

3. Branger C, O’Connell ME and Morgan DG. Factor Analysis of the 12-Item Zarit Burden Interview in Caregivers of Persons Diagnosed With Dementia. J Appl Gerontol 2016; 35: 489–507. 20140130. DOI: 10.1177/0733464813520222.

4. Novak M and Guest C. Application of a multidimensional caregiver burden inventory. Gerontologist 1989; 29: 798–803. DOI: 10.1093/geront/29.6.798.

5. Sharif Nia H, Hosseini L, Ashghali Farahani M, et al. Development and validation of care stress management scale in family caregivers for people with Alzheimer: a sequential-exploratory mixed-method study. BMC Geriatr 2023; 23: 82. 20230207. DOI: 10.1186/s12877-023-03785-6.

6. Pitsikali A, Galanakis M, Varvogli L, et al. Kingston Caregiver Stress Scale (KCSS) Greek Validation on Dementia Caregiver Sample. Psychology 2015; 6: 1180–1186. DOI: 10.4236/psych.2015.69116.

7. Wuttke-Linnemann A, Palm S, Scholz L, et al. Introduction and Psychometric Validation of the Resilience and Strain Questionnaire (ResQ-Care)-A Scale on the Ratio of Informal Caregivers’ Resilience and Stress Factors. Front Psychiatry 2021; 12: 778633. 20211124. DOI: 10.3389/fpsyt.2021.778633.

8. Seng BK, Luo N, Ng WY, et al. Validity and reliability of the Zarit Burden Interview in assessing caregiving burden. Ann Acad Med Singap 2010; 39: 758–763.

9. Le Toullec E, Le Gagne A, Leblong E, et al. Assessment of burden and needs of family caregivers for the elderly, a scoping review. Front Aging 2025; 6: 1578911. 20250613. DOI: 10.3389/fragi.2025.1578911.

10. Ismail Z, Guan DX, Vellone D, et al. The Canadian platform for research online to investigate health, quality of life, cognition, behaviour, function, and caregiving in aging (CAN-PROTECT): Study protocol, platform description, and preliminary analyses. Aging and Health Research 2024; 4: 100207.

11. Mudalige D, Guan DX, Ballard C, et al. The mind and motion: exploring the interplay between physical activity and Mild Behavioral Impairment in dementia-free older adults. Int Rev Psychiatry 2024; 36: 196–207. 20240622. DOI: 10.1080/09540261.2024.2360561.

12. Warring I, Guan D, Ballard C, et al. Mild Behavioral Impairment and Quality of Life in Community Dwelling Older Adults. Int J Geriatr Psychiatry 2024; 39: e6153. DOI: 10.1002/gps.6153.

13. Crockford JFE, Guan DX, Einstein G, et al. Menopausal symptom burden as a predictor of mid-to late-life cognitive function and mild behavioral impairment symptoms: A CAN-PROTECT study. PLoS One 2025; 20: e0301165. 20250305. DOI: 10.1371/journal.pone.0301165.

14. Guan DX, Mortby ME, Pike GB, et al. Linking cognitive and behavioral reserve: Evidence from the CAN-PROTECT study. Alzheimers Dement (N Y) 2024; 10: e12497. 20241004. DOI: 10.1002/trc2.12497.

15. Guan DX, Aundhakar A, Tomaszewski Farias S, et al. Vascular risk factor associations with subjective cognitive decline and mild behavioural impairment. Brain Commun 2025; 7: fcaf163. 20250428. DOI: 10.1093/braincomms/fcaf163.

16. Phelps J, Guan DX, Teo K, et al. Validation of the standard assessment of global everyday activities (SAGEA) scale for dementia diagnosis. Age and Ageing 2025; 54. DOI: 10.1093/ageing/afaf115.

17. Miller DS, Robert P, Ereshefsky L, et al. Diagnostic criteria for apathy in neurocognitive disorders. Alzheimers Dement 2021; 17: 1892–1904. 20210505. DOI: 10.1002/alz.12358.

18. Sano M, Cummings J, Auer S, et al. Agitation in cognitive disorders: Progress in the International Psychogeriatric Association consensus clinical and research definition. Int Psychogeriatr 2024; 36: 238–250. 20230307. DOI: 10.1017/S1041610222001041.

19. Cummings J, Pinto LC, Cruz M, et al. Criteria for Psychosis in Major and Mild Neurocognitive Disorders: International Psychogeriatric Association (IPA) Consensus Clinical and Research Definition. Am J Geriatr Psychiatry 2020; 28: 1256–1269. 20200905. DOI: 10.1016/j.jagp.2020.09.002.

20. Fischer CE, Ismail Z, Youakim JM, et al. Revisiting Criteria for Psychosis in Alzheimer’s Disease and Related Dementias: Toward Better Phenotypic Classification and Biomarker Research. J Alzheimers Dis 2020; 73: 1143–1156. DOI: 10.3233/JAD-190828.

21. Ismail Z, Smith EE, Geda Y, et al. Neuropsychiatric symptoms as early manifestations of emergent dementia: Provisional diagnostic criteria for mild behavioral impairment. Alzheimers Dement 2016; 12: 195–202. 20150618. DOI: 10.1016/j.jalz.2015.05.017.

22. Kroenke K, Spitzer RL and Williams JB. The PHQ-9: validity of a brief depression severity measure. J Gen Intern Med 2001; 16: 606–613. DOI: 10.1046/j.1525-1497.2001.016009606.x.

23. Spitzer RL, Kroenke K, Williams JB, et al. A brief measure for assessing generalized anxiety disorder: the GAD-7. Arch Intern Med 2006; 166: 1092–1097. DOI: 10.1001/archinte.166.10.1092.

24. Hughes ME, Waite LJ, Hawkley LC, et al. A Short Scale for Measuring Loneliness in Large Surveys: Results From Two Population-Based Studies. Res Aging 2004; 26: 655–672. DOI: 10.1177/0164027504268574.

25. Thase ME, Ismail Z, Meehan SR, et al. Assessment of patient life engagement in major depressive disorder using items from the Inventory of Depressive Symptomatology Self-Report (IDS-SR). J Psychiatr Res 2023; 161: 132–139. 20230206. DOI: 10.1016/j.jpsychires.2023.02.008.

26. Guttman L. A basis for analyzing test-retest reliability. Psychometrika 1945; 10: 255–282. DOI: 10.1007/bf02288892.

27. Gosling CJ, Colle R, Cartigny A, et al. Measuring loneliness: a head-to-head psychometric comparison of the 3-and 20-item UCLA Loneliness Scales. Psychol Med 2024; 54: 1–7. 20241031. DOI: 10.1017/s0033291724002083.

28. Ismail Z, Warring I, Guan DX, et al. Validation of the Quality of Life and Function Five Domain Scale (QFS-5) against the EuroQol-5D (EQ-5D). medRxiv 2025: 2025.2008.2031.25334811. DOI: 10.1101/2025.08.31.25334811.

29. Weiss C, Meehan SR, Brown TM, et al. Ejects of adjunctive brexpiprazole on calmness and life engagement in major depressive disorder: post hoc analysis of patient-reported outcomes from clinical trial exit interviews. J Patient Rep Outcomes 2021; 5: 128. 20211211. DOI: 10.1186/s41687-021-00380-4.

30. Terwee CB, Bot SD, de Boer MR, et al. Quality criteria were proposed for measurement properties of health status questionnaires. Journal of clinical epidemiology 2007; 60: 34–42.

31. McHorney CA and Tarlov AR. Individual-patient monitoring in clinical practice: are available health status surveys adequate? Quality of life research 1995; 4: 293–307.

32. Rodenberg CA. A Review of: “Health Measurement Scales: A Practical Guide to Their Development and Use, Fourth Edition, by D. L. Streiner and G. R. Norman”. Journal of Biopharmaceutical Statistics 2009; 19: 1162–1164.

33. Cohen J. Statistical Power Analysis for the Behavioral Sciences. 2nd ed. Hillsdale, N.J.: L. Erlbaum Associates, 1988.

34. Carmines EG and McIver C. Analyzing models with unobserved variables: analysis of covariance structures. In: 1981.

35. Clark LA and Watson D. Constructing validity: Basic issues in objective scale development. Psychological Assessment 1995; 7: 309–319.

36. Gallagher D, Ni Mhaolain A, Crosby L, et al. Dependence and caregiver burden in Alzheimer’s disease and mild cognitive impairment. Am J Alzheimers Dis Other Demen 2011; 26: 110–114. 20110113. DOI: 10.1177/1533317510394649.

37. Denham AMJ, Wynne O, Baker AL, et al. An online survey of informal caregivers’ unmet needs and associated factors. PLoS One 2020; 15: e0243502. 20201210. DOI: 10.1371/journal.pone.0243502.

38. Cheng ST. Dementia Caregiver Burden: a Research Update and Critical Analysis. Curr Psychiatry Rep 2017; 19: 64. 20170810. DOI: 10.1007/s11920-017-0818-2.

39. Cheng ST, Au A, Losada A, et al. Psychological Interventions for Dementia Caregivers: What We Have Achieved, What We Have Learned. Curr Psychiatry Rep 2019; 21: 59. 20190606. DOI: 10.1007/s11920-019-1045-9.

40. Schulz R and Sherwood PR. Physical and mental health ejects of family caregiving. Am J Nurs 2008; 108: 23–27; quiz 27. DOI: 10.1097/01.NAJ.0000336406.45248.4c.

41. Pinquart M and Sörensen S. Dijerences between caregivers and noncaregivers in psychological health and physical health: a meta-analysis. Psychol Aging 2003; 18: 250–267. DOI: 10.1037/0882-7974.18.2.250.

42. Mahoney R, Regan C, Katona C, et al. Anxiety and depression in family caregivers of people with Alzheimer disease: the LASER-AD study. Am J Geriatr Psychiatry 2005; 13: 795–801. DOI: 10.1176/appi.ajgp.13.9.795.

43. Sheehan OC, Haley WE, Howard VJ, et al. Stress, Burden, and Well-Being in Dementia and Nondementia Caregivers: Insights From the Caregiving Transitions Study. Gerontologist 2021; 61: 670–679. DOI: 10.1093/geront/gnaa108.

44. Pinquart M and Sörensen S. Gender dijerences in caregiver stressors, social resources, and health: an updated meta-analysis. J Gerontol B Psychol Sci Soc Sci 2006; 61: P33–45. DOI: 10.1093/geronb/61.1.p33.

45. World Health Organization. Global action plan on the public health response to dementia 2017-2025. 2017. Geneva, Switzerland.

46. Oh E, Moon S, Chung D, et al. The moderating eject of care time on care-related characteristics and caregiver burden: dijerences between formal and informal caregivers of dependent older adults. Front Public Health 2024; 12: 1354263. 20240404. DOI: 10.3389/fpubh.2024.1354263.

