## Supplementary Figure 1 for "Validation of the Care Partner Stress Scale in the CAN-PROTECT Study"

**For all the questions below, please answer the following question and score each from 1-7.**

With respect to being responsible for, or caring for a person with a cognitive or neurological disorder **in the last month,** how often did any of the following contribute to your level of stress?

| 1 | 2 | 3 | 4 | 5 | 6 | 7 |
|---|---|---|---|---|---|---|
| never | rarely | occasionally | sometimes | often | frequently | constantly |

| **Cognitive Decline** | **Score** |
|---|---|
| 1. Forgetfulness or difficulties learning (e.g., having to repeat yourself, hearing repetitive stories or instructions, looking for lost items, teaching how to use technology, collectively making decisions based on conversations which are later forgotten, dealing with factual inconsistencies or made-up conversations) | _____ |
| 2. Inattention (e.g., unable to focus, pay attention, or stay alert) | _____ |
| 3. Difficulties with communication due to decline in language abilities (understanding or making yourself understood) | _____ |
| 4. Recognition (e.g., not recognizing places and easily getting lost or wandering off, not recognizing familiar people) | _____ |
| 5. Decision making (e.g., poor planning and organization) | _____ |
| **Behaviour Changes** | |
| 1. Apathy (loss of interest, drive, motivation, or emotional reactivity) | _____ |
| 2. Emotional symptoms (low mood, anxiety, worry) | _____ |
| 3. Excessive motor activity (e.g., pacing, rummaging, trying to leave) | _____ |
| 4. Verbal aggression (e.g., being argumentative, having a bad temper, poor frustration tolerance, verbal outbursts, rudeness) | _____ |
| 5. Physical aggression (e.g., resisting care, grabbing, pushing, hitting, scratching) | _____ |
| 6. Hoarding or repetitive behaviours | |
| 7. Disinhibited or socially inappropriate behaviour (e.g., loss of tact, empathy, or social graces, disclosing personal or intimate details, intruding on others, sexually inappropriate behaviour, or uncomfortable discussions about sex) | _____ |
| 8. Psychotic symptoms (e.g., suspiciousness, paranoid thinking, seeing/hearing things) | _____ |
| **Functional Impairment** | |
| 1. Assisting with activities of daily living (bathing, dressing, eating, transferring, toileting, incontinence) | _____ |

© Zahinoor Ismail MD 2021

2. Managing instrumental activities of daily living (e.g., finances, transportation, shopping and/or meal preparation, housecleaning and/or home maintenance, managing communication (telephone, mail, email), medication management) ______
3. Attending or assisting with medical appointments ______
4. Dealing with legal issues ______
5. Overwhelming number of tasks involved in caregiving ______
6. Amount of time every week spent on caregiving obligations ______
7. Loss of insight into changes in function (or cognition or behaviour) ______

**Unmet needs and emotional impact on caregiver**

1. Not having your own needs met ______
2. Not enough time for yourself or your interests ______
3. Not being able to take care of yourself ______
4. Limited social life ______
5. Feeling like you are missing out on other things ______
6. Feelings of guilt or inadequacy to provide care ______

**Work interference/ financial strain**

1. Missing work or poorer work quality due to caregiving obligations ______
2. Financial hardship related to current caregiving ______
3. Concerns over how to pay for future caregiving needs ______
4. Safeguarding the person from financial exploitation or scams ______

**Family interference/ interpersonal conflict**

1. Detrimental effect of caregiving on your own personal relationships ______
2. Not having enough time for your own family ______
3. Conflict with family over how to provide care ______
4. Conflict with family over time spent caregiving ______
5. Feeling underappreciated by family members for efforts in providing care ______

**Situational Perception**

1. Feeling unable to control the situation ______
2. Not knowing enough about the disease or condition ______
3. Being unprepared for new roles/responsibilities (e.g., managing finances) ______
4. Feeling like this responsibility should not be yours ______
5. Feeling stuck or trapped in this role of caregiver ______
6. Not seeing an end to caregiving ______

© Zahinoor Ismail MD 2021

© 2021 ZAHINOOR ISMAIL MD

| | |
|---|---|
| 7. Not having enough family support or help for caregiving duties | ______ |
| 8. Not having enough formal supports (e.g., home care, day program, or respite care) | ______ |
| 9. Not feeling on top of things because of caregiving obligations | ______ |
| 10. Having to be responsible due to gender role expectations | ______ |
| 11. Cultural expectations to take on the caregiver role | ______ |

© Zahinoor Ismail MD 2021
